# Identification of genetic variants in Pfs25 and functional evaluation in mosquito infection

**DOI:** 10.64898/2026.08.25.26361130

**Authors:** Alessandra Orfano, Awa Cisse, Yicheng Guo, Leeah Han, Nathan Fikadu, Laty G. Thiam, Aboubacar Ba, Rebecca Li, Mariama N. Pouye, Khadidiatou Mangou, Adam J. Moore, Fatoumata Diallo, Seynabou D. Sene, Elhadji Malick Ngom, Bacary D. Sadio, Alassane Mbengue, Christopher Membi, Billy Ngasala, Thomas Bazie, Fabrice Anyirékun Some, Natalie Olson, Saurabh D. Patel, Lawrence Shapiro, Sunil Parikh, Brian D. Foy, Michael Cappello, Ines Vigan-Womas, Zul Premji, Roch K. Dabire, Jean-Bosco Ouedraogo, Zizhang Sheng, Amy K. Bei

## Abstract

Transmission-blocking vaccines (TBVs) are a promising strategy to reduce malaria transmission by targeting parasite stages within the mosquito. However, parasite genetic diversity may limit vaccine efficacy. We used next-generation amplicon deep sequencing to identify non-synonymous single nucleotide polymorphisms (SNPs) in Pfs25 from 184 *Plasmodium falciparum* isolates from Senegal, Tanzania, Ghana, and Burkina Faso. Prioritized SNPs were introduced into *P. falciparum* via CRISPR-Cas9. For the G116C variant, gametocyte development was evaluated by microscopy and qPCR, and mosquito infectivity was assessed by SMFAs. We identified 26 SNPs, including 24 novel variants. Functional assays showed that the Pfs25 G116C mutation did not affect gametocyte development or exflagellation. SMFA showed no significant differences in oocyst prevalence or intensity between mutant and WT parasites. These findings highlight the importance of integrating genetic surveillance with functional validation to guide the development of effective transmission blocking interventions.

## Introduction

Malaria is a deadly disease with a global impact marked by the co-evolution of *Plasmodium* spp. and the Anopheles mosquito vector. In 2024 alone, an estimated 282 million cases of malaria were reported, resulting in over 600 thousand deaths^1^. Over 90% of these deaths occurred in sub-Saharan Africa, with a significant burden in children and pregnant women, causing a massive health and economic burden for affected countries. Despite some malaria cases being treatable, the emergence of drug-resistant parasites and insecticide-resistant mosquitoes have posed enormous threats to malaria control, and these challenges have contributed to stagnation, and in some regions, an increase in malaria infections^2–6^.

Recently, following decades of development and the successful completion of Phase III clinical trials^7,8^ the World Health Organization has recommended two pre-erythrocytic malaria vaccines, RTS,S/AS01 and R21/Matrix-M, to prevent malaria infections caused by *P. falciparum* in children living in high-risk areas. This is a remarkable advancement in the fight against malaria. However, antigenic diversity remains a critical factor for acquiring highly protective immunity and a major barrier to developing an effective next-generation vaccine. To date, the most advanced malaria vaccines have shown only modest protective efficacy^9,10^.

Among the many different strategies targeting disease vectors or the pathogens they transmit^11–14^ such as insecticide-treated bed nets, indoor residual spraying, genetically modified mosquitoes and transmission-blocking vaccines (TBVs) represent a promising approach as the goal shifts to malaria elimination^15,16^. TBVs induce antibodies that target the Plasmodium antigens expressed in sexual stages such as gametocyte (Pfs230, Pfs48/45), gamete (Pfs230, Pfs48/45), zygote (Pfs25), and ookinete (Pfs25, Pfs28), disrupting the parasite life cycle inside the mosquito and preventing transmission to the human instead of directly preventing infection or clinical symptoms within the human host. This results in indirect community protection rather than individual direct protection^17^.

Pfs25 is a *P. falciparum* sexual stage cysteine-rich protein displayed on the surface of zygotes and ookinetes, which are present in the mosquito midgut upon fertilization^18,19^. Pfs25 has a predicted role in parasite survival in the protease-rich mosquito midgut and in crossing the midgut epithelium^20,21^. With success in generating a recombinant Pfs25 protein and preclinical evidence of superior functional serum activity^22–24^, Pfs25 and its orthologue Pvs25 have been prioritized for TBV clinical development^25–33^. The Pfs25 antigen incorporated into various vaccine delivery platforms has been evaluated extensively in clinical studies for the transmission-blocking activity and potently inhibits oocyst development. Still, there is a rapid decline in antibody titers *in vivo* post-vaccination after four doses^29–32^. Since the Pfs25 protein is only present on gametes, zygotes, and ookinetes in the mosquito, the Pfs25 TBV cannot rely on natural boosting to increase antibody titers after vaccination in the human. Also, a new study characterizes additional transmission-reducing epitopes for a more potent Pfs25 TBV^34^. Although recent clinical evidence has demonstrated that Pfs230 elicits stronger and more durable transmission blocking activity than Pfs25^35^, Pfs25 remains an attractive vaccine target because of its high sequence conservation across geographically diverse populations. Consequently, multivalent vaccine strategies combining Pfs25 with another antigen, possibly Pfs230, have shown promising results in mouse models using the mRNA vaccine platform for TBV^36^.

One of the major challenges that limits the ability to formulate an efficacious vaccine is the vast antigenic diversity of the parasite, which plays a significant role in immune evasion. Consequently, the impact of the biological interactions with the mosquito vector compromises the development of potential vaccines or even the effectiveness of those already in trial^18,19,36,37^. Understanding the genetic diversity of Pfs25 is essential for rational and structure-guided design of effective TBVs. Genetic variations within Pfs25 can potentially influence the efficacy of these vaccines, potentially affecting their ability to interrupt the transmission cycle. There are limited data on the impact of *Plasmodium* population genetic diversity of vaccine candidates like Pfs25^37–40^ and studies addressing the genetic diversity and discovery of novel SNPs are needed to investigate the significant impact of these novel SNPs on transmission and immune evasion. In this study, we investigate the genetic diversity of Pfs25 in complex infections from four highly endemic countries across Africa: Burkina Faso, Ghana, Senegal, and Tanzania, and functionally evaluate selected variants for their impact on mosquito infection using genome-edited parasites.

## Materials and Methods

### Ethics statement

Samples used in this study are archived samples collected in endemic regions with complex *P. falciparum* infections. All samples from this study received approval from both the Human Investigation Committee of Yale University and the local ethical review committees: Tanzania: Commission for Science and Technology (Permit No. 2003-207-CC-2003-102) and Harvard School of Public Health Human Subjects Committee (HSC Protocol #P11778-101); Senegal: National Ethics Committee of Senegal (CNERS) (SEN19/36), the regulatory board of the Senegalese Ministry of Health and the Institutional Review Board of the Yale School of Public Health (2000025417), Burkina Faso: Human Investigation Committee of Yale University (IRB Protocol 2000024009) and Comite D’Ethique Institutionnel pour la Recherche en Sciences de la Sante (A009-2018), Ghana: approved by the Yale University Human Investigations Committee (IRB protocol: 2000022860). All research was performed in accordance with relevant guidelines and regulations, and informed consent was obtained from all participants and/or their legal guardians.

Human blood used for mosquito feeding and parasite cultures was obtained under a protocol approved by the Yale University Human Investigation Committee (IRB/HIC protocol # 2000034487). Consent was obtained from all blood donors. All mosquito rearing and infection experiments were performed in accordance with institutional and federal guidelines for arthropod-based research.

### Mosquito rearing

*Anopheles gambiae* (G3 strain) mosquitoes were reared in the insectary at the Yale School of Public Health. Larvae were maintained in trays with picotap water, at 26 °C under a 12h light/dark cycle and fed ground fish food. Adult mosquitoes were kept at 26 °C and 70% relative humidity with a 12h light/dark cycle. A 10% corn syrup solution provided on cotton pads was used as the sugar source. For egg production, mosquitoes were fed O+ human blood obtained under an IRB-approved protocol 2000034487.

### Study sites

Samples used in this study are archived *P. falciparum* positive samples collected from malaria endemic regions with complex infections across Burkina Faso, Ghana, Senegal, and Tanzania (Fig.1). Detailed descriptions of each site are provided below:

**Figure 1:**
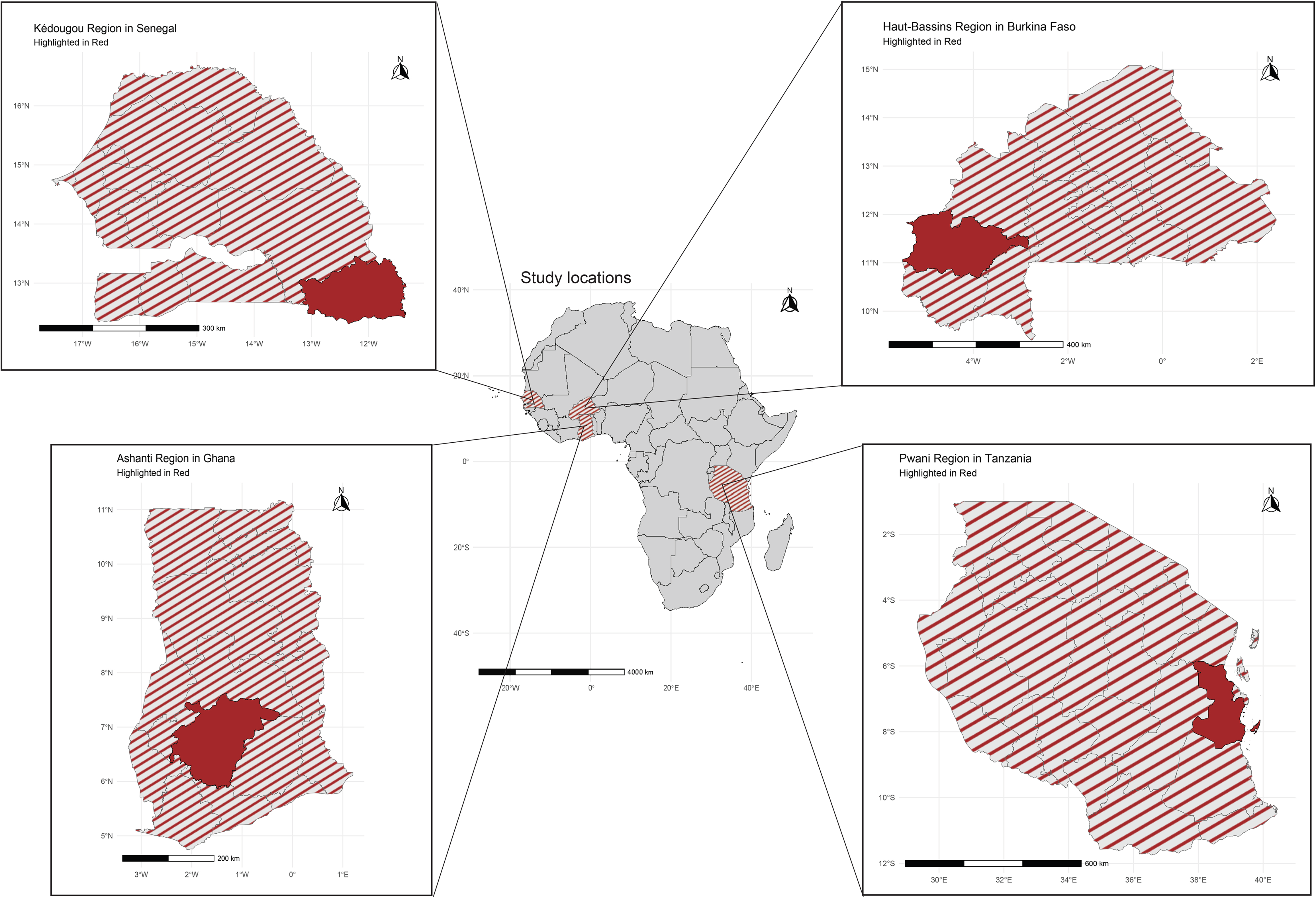
Geographic locations of study sites in Africa. Highlighted in red are the Kédougou Region (Senegal), Haut-Bassins Region (Burkina Faso), Ashanti Region (Ghana), and Pwani Region (Tanzania). The central map of Africa displays the locations of the four countries where study sites are situated, with expanded panels highlighting the specific regions included in the analysis. Study regions were selected to capture diverse malaria transmission settings. The entomological inoculation rate (EIR), a measure of malaria transmission intensity, varies across sites: Kédougou, Senegal (EIR ≈ 250); Bama, Burkina Faso (EIR ≈ 200); Kumasi, Ghana (EIR ≈ 193); and Pwani, Tanzania (EIR ≈ 367). Maps were created in Rstudio.

#### Burkina Faso

Samples were collected in 2018 in Bama, a rural area located in Dandé in the Kou Valley. The region is highly endemic and experiences a rainy season from June to October. The annual entomological inoculation rate (EIR) reaches up to 200 infective bites/person/night^41^. The predominant vector is *An. gambiae*, with *An. funestus* present in lower proportions.

#### Ghana

Samples were collected in 2018 from six schools (Darko Primary, Fankyenebra K-8, Martyrs of Uganda Prep, Opoku Ware Primary, and Darko JHS) in Kumasi, Ghana. Malaria transmission peaks between July and November, with *An. gambiae* as the predominant vector. The reported annual EIR is up to 193 infective bites/ person/night^42^.

#### Senegal

Samples were collected in 2019 and 2022 as part of ongoing malaria surveillance led by the Institute Pasteur de Dakar, from six health posts in Kédougou: Camp Militaire, Bandafassi, Bantako, Mako, Tomboronkoto and Dalaba. The rainy season spans May to November, with peak malaria transmission during this period. The predominant malaria vector species is *An. gambiae* and *An. funestus* and the annual EIR is approximately 250 infective bites/ person/night^43^.

#### Tanzania

Samples were collected in 2004 from the Mlandizi Health Centre in the Kibaha district of the Pwani coastal region, located about 40 km northwest of Dar es Salaam. Malaria transmission is perennial with seasonal peaks from May – July and December-January. The main malaria vector species are members of the *An. gambiae* complex and *An. funestus*, with a mean annual EIR of 367 infective bites/ person/night, ranging from 94-667^44^.

### DNA extraction

DNA was extracted from dried blood spots (DBS) using the QIAmp DNA Blood mini kit (Qiagen) following the manufacturer’s instructions for DBS extractions. Briefly, three 3 mm punches were taken from each DBS sample and placed into 1.5 ml microcentrifuge tubes. After extraction, column binding, and washing, DNA was eluted in two successive 35 μL elutions from the same column (representing E1 and E2) into sterile tubes and stored at -20°C.

### Multiplicity of infection (MOI)

The number of distinct *P. falciparum* genotypes in each sample was determined based on polymorphisms in the merozoite surface protein -1 (MSP-1) and MSP-2 genes. The MSP genotyping of the *P. falciparum* isolates was performed using nested PCR as described by Snounou^45^. Briefly, the primary PCR reaction consisted of 12.5 μL of GoTaq Green Master Mix (Promega) (containing Taq DNA polymerase, dNTPs, MgCl_2,_ and reaction buffer), 0.125 μM of each primer, 10.9 μL of nuclease-free water, and 1 μL of genomic DNA. For the nested PCR, 1 μL of the first-round PCR product was used as a template. The nested PCR products were analyzed by size differences using 3% agarose gel electrophoresis.

Allelic families targeted for genotyping included K1, MAD20, and RO33 for MSP1 and FC27 and IC for MSP2, as previously described^45^. The number of genetically distinct clones per isolate was defined as the highest number of alleles detected in either MSP1 or MSP2. The average MOI for each population was calculated by summing the total number of clones detected across all isolates and dividing by the number of samples analyzed.

### Polymerase chain reaction (PCR) and sequencing of the Pfs25 gene

Primers were designed to amplify the full length of the Pfs25 gene (654 bp): forward primer (Pfs25 F): 5’-ATGAATAAACTTTACAGTTTGTTTC-3’ and reverse primer (Pfs25 R): 5’-TTACATTATAAAAAAGCATACTG-3’. PCR was performed using the Platinum Taq DNA Polymerase High-Fidelity (Invitrogen) which has proofreading capability. The reaction consisted of 10X High Fidelity PCR Buffer, 10 μM forward primer,10 μM reverse primer, 10 μM deoxynucleotide triphosphates (dNTPs), 0.2 μL Platinum Taq DNA Polymerase Hi-Fidelity Enzyme, and 2 μL DNA template in a 25-μL volume. The thermocycling conditions were initial denaturation at 94°C for 5 minutes, followed by 39 cycles at 94°C for 30 seconds, 58°C for 30 seconds, and 68°C for 1 minute, and a final extension at 68°C for 5 minutes. Amplicon size was confirmed by agarose gel electrophoresis before proceeding with library preparation for Next Generation Sequencing (NGS).

The library prep of the samples was performed as described by Mangou et al^46^. Briefly, PCR amplicons were bead purified using Omega magnetic beads and quantified using a Qubit fluorometer. Concentrations were normalized prior to library preparation with the Nextera XT DNA Library Prep kit (Illumina). Unique dual indexes (UDIs) were added to each sample, followed by a second bead purification and quantification using the KAPA library Quantification kit (Roche). Libraries were normalized to a concentration of 4nM, pooled into eight sub-pools, bead purified again, quantified, and combined in equal volumes to generate a final pool. Sequencing was performed at the Yale Center for Genome Analysis (YCGA) on the NovaSeq 6000 platform, targeting approximately 500,000 reads per sample. Of the 201 clinical isolates sequenced, 184 yielded sufficient sequence data for analysis and were included in the study.

### Sequence assembly and polymorphism analysis

De-multiplexed forward and reverse reads for all samples were obtained from the Yale Center for Genome Analysis (YCGA). Using Geneious Prime, sequences were paired and trimmed with the BBDuk plugin, applying a minimum quality score of 30 and a minimum read length of 75bp. Trimmed reads were mapped to the Pfs25 reference gene from the P. falciparum 3D7 strain (PF3D7_1031000). Non-synonymous single nucleotide polymorphisms (SNPs) were called using a minimum variant frequency threshold of 1% (0.01) and a minimum coverage of 500 reads per site. To ensure accuracy and reproducibility, at least three individuals independently performed sequence alignment and SNP calling for each sample. As a control for PCR and sequencing fidelity, Pfs25 was also amplified and sequenced from genomic DNA of the 3D7 reference, NF54, and Dd2 strains alongside each batch of test samples. These controls helped verify that rare SNPs were not artifacts of PCR or sequencing errors when applying variant frequency threshold of 1%.

### Protein structure and prediction of SNP function

Individual FASTA files were generated for the amino acid sequences of Pfs25 and its variants containing the identified SNPs. These sequences were threaded onto the crystal structure of the Pfs25, and PyMol (version 2.3.2) was used to visualize the structural location of each SNP and to assess their potential impact. The effects of each SNP were evaluated in the context of known inhibitory antibody binding sites to predict potential roles in immune evasion. Structural alterations caused by the mutations were assessed, and binding affinity between mutant versions of the protein and Pfs25 was estimated. FoldX was used to calculate the change in binding free energy (delta delta G) associated with each SNP, providing an estimate of their impact on protein stability and interactions (Supplementary table S1).

### Plasmid cloning of Pfs25 allelic replacement constructs for CRISPR-Cas9 editing

The plasmid pDC2-cam-Cas9-U6-sgRNA-hDHFR, optimized for *P. falciparum*, was used to generate the single-vector Crispr-Cas9 constructs. This plasmid expresses Streptococcus pyogenes Cas9 (SpCas9) under the control of the calmodulin promoter, a single guide RNA (sgRNA) under the control of the U6 promoter, and includes the selectable marker hDHFR^47,48^. Guide RNAs were selected based on high scores and minimal off-target effects, within a region spanning 100-200bp from the point mutation. The gRNAs were synthesized as oligonucleotides with the Bbs I compatible overhangs, in addition to a 5’ G upstream of the N20 of the guide. The pDC2 vector was digested with Bpil, and annealed oligos were phosphorylated and ligated into the digested vector.

The two homology regions (donor DNAs) were amplified by PCR from *P. falciparum* NF54 genomic DNA using specific primers B1 with Pfs25 wt synthetic R and A4 with pfs25 Synthetic wt F (Supplementary table S2). A segment of the donor sequence encompassing the edit site and gRNA binding site was recodonized (the nucleotide sequence was altered without changing the amino acid sequence) to prevent CRISPR/Cas9 system from cutting the plasmid, permitting only cutting of the target gene in the genome. The recodonized region was synthesized and PCR amplified.

The full donor DNA construct was assembled by Gibson assembly into the digested pDC2 vector. The construct containing the point mutation was created from two PCR fragments, the mutation was introduced via primer-directed mutagenesis, using the sequence confirmed WT-replacement donor plasmid as a template. The two fragments were assembled into the pDC2 backbone and transformed into *E. coli* competent cells. Individual colonies were selected, plasmid purified, and insert region was sequence-confirmed by Sanger sequencing. Final plasmids were further confirmed by full-plasmid sequencing (Plasmidsaurus).

### Transfection of *P. falciparum*

Asexual parasites of the NF54 line (MRA-1000, line E) were obtained from the BEI Resources Repository. A total of 100ng of plasmid DNA was ethanol precipitated, resuspended in cytomix, and used to transfect NF54 ring stage parasites at 5% parasitemia using the Biorad Gene Pulser electroporator. Transfected cells were transferred to a culture plate containing 10ml of complete RPMI medium and 200 μL of O^+^ human red blood cells (RBCs). Twenty-four hours post-transfection, blood smears were prepared to assess viable parasitemia, and selection with 2.5nM WR99210 was performed for five consecutive days. When bulk transfected cultures became microscopy positive, blood pellets were collected for genomic DNA extraction to confirm genomic integration by PCR and Sanger sequencing. Cloning by limiting dilution was performed to isolate multiple individual clonal lines, which were further verified to contain the edited SNP of interest by PCR and Sanger sequencing.

### Gametocyte induction

Two clones of mutant parasite NF54^G116C^ (clones 5B and 8E) and wild–type NF54 were cultured asexually and gametocytogenesis was induced as previously described^49^. The inductions were carried out in 6-well plates, with cultures adjusted to 2% parasitemia and 5% hematocrit on day 1, in the absence of drug. Media was replaced daily throughout the induction period, during which mature gametocytes (stages IV and V) were obtained. Samples for RNA extraction were collected in duplicate on days 4, 6, 8, 10, 12, and 13 and preserved with TriReagent. Prior to sample collection, thin blood smears were prepared to quantify gametocyte stage conversion by microscopy.

On day 13 post-induction, 20 µL of each parasite culture was collected and centrifuged to remove the medium. The pellet was resuspended in an equal volume of O+ human serum, thoroughly mixed, and then mounted on a slide with a coverslip. Exflagellation centers were counted in ten randomly selected microscopic fields at 40x magnification, and the mean number per field was calculated. Stage V gametocytemia was determined separately using a Giemsa-stained thin smear prepared from the same culture.

For mosquito feeding, cultures were adjusted to final gametocytemias of 0.5% or 0.2% at 40% hematocrit in fresh O+ human serum and O+ erythrocytes. All cultures were standardized to the same gametocytemia before feeding. *An. gambiae* G3 mosquitoes, 5–7 days old, were used for infection. Following feeding, mosquitoes were maintained at 26 °C and 70% relative humidity and provided with 10% sugar solution (corn syrup) until the dissection of the midgut.

### RNA Extraction, DNAse Digest and Reverse Transcription

RNA from gametocyte cultures, collected in duplicate and stored in TriReagent (Invitrogen), was isolated following the manufacturer’s protocol with minor modifications. Briefly, chloroform was added, samples were centrifuged at 4 °C, and the aqueous phase was transferred to a new tube. An equal volume of ethanol containing β-mercaptoethanol (BME) was added, and samples were loaded onto PureLink RNA Mini Kit columns (Invitrogen, PureLink) and proceeded to step 3 of the manufacturer’s protocol. RNA was precipitated overnight at –20 °C with 100% ethanol, 3M sodium acetate, and glycogen blue. Pellet was washed with 70% ethanol and resuspended in RNase-free water at a final concentration of 100 ng/µL. cDNA was synthesized using the SuperScript IV First-Strand Synthesis Kit (Invitrogen) with ezDNase (Invitrogen) treatment.

### Quantitative Real-Time PCR

For gene expression analysis, primers were used to detect transcripts representative of different stages of gametocyte development: PF14_0748 (early), Pfs48/45 (mid), and Pfs25 (late), as described previously^50^. The ubiquitin conjugation enzyme (PF08_0085) was used as the constitutive control. However, the Pfs25 primer set was redesigned because the original primers described by Joice et al^50^. targeted a region that had been recodonized in the transgenic construct, preventing their use for transcript detection.

Quantitative RT-PCR was performed using diluted cDNA and run on a CFX96 Bio-Rad Real-Time PCR system with the following program: initial denaturation at 95 °C for 10 min; 40 cycles of 95 °C denaturation for 30s, 58 °C annealing for 1 min, and a final dissociation protocol from 55 °C to 95 °C with 0.5 °C increments every 30 s to verify primer specificity and product formation. Each reaction included a negative control (–RT) to exclude genomic DNA contamination. All reactions were performed in triplicate, and mean threshold cycle (Ct) values were used for analysis.

## Results

### Characteristics of study participants

The 184 samples used in this study were obtained from individuals between the ages of 1 to 74 years, but many of them ranged from 1-19 years. Samples were collected in 2004 and between 2018 and 2022 in four regions in sub-Saharan Africa: Kumasi (Ghana), Kédougou (Senegal), Pwani (Tanzania), and Bama in Haut-Bassins region (Burkina Faso) (Fig. 1). Additional demographic information is summarized in Table 1.

**Table 1.** Summary of patient demographics and infection complexity across study sites. The table shows the number of patients recruited from each country (Burkina Faso, Ghana, Senegal, and Tanzania) and the total. The sex ratio is expressed as the proportion of male to female participants. The complexity of infection (COI) is reported as the mean for each country. * Statistical differences were calculated using the Chi-square test. ** Statistical differences were calculated using Kruskal-Wallis test.

| Countries | Burkina Faso | Ghana | Senegal | Tanzania | Total | p- value |
| --- | --- | --- | --- | --- | --- | --- |
| **Participants (number)** | 31 | 55 | 73 | 25 | 184 |  |
| **Sex** |  |  |  |  |  |  |
| **Male** | 12 | 27 | 31 | 8 | 78 |  |
| **Female** | 22 | 28 | 42 | 17 | 108 |  |
| **Unknown** | 4 | 1 | 2 | 0 | 8 |  |
| **Sex ratio (male/female)** | 0.57 | 0.96 | 0.73 | 0.47 | 0.76 | 0.424* (ns) |
| **Age (yr) (min- max)** | 1.5-32 | 5-16 | 2-74 | 1-52 |  |  |
| **<9** | 20 | 20 | 12 | 16 |  |  |
| **10-19** | 11 | 36 | 30 | 4 |  |  |
| **20-30** | 2 | 0 | 20 | 2 |  |  |
| **31-50** | 1 | 0 | 9 | 0 |  |  |
| **>51** | 0 | 0 | 2 | 1 |  |  |
| **Unknown** | 4 | 0 | 2 | 2 |  |  |
| **COI/MOI (mean)** | 2.27 | 2.51 | 4.03 | 2.1 |  | p < 0.001** |

In Senegal, samples were collected in July 2019 and 2022 from five clinics in Kédougou, Senegal through passive case detection. The criteria eligibility for the study is the temperature equal to or greater than 38°C or a fever in the past 24 hours and positive for malaria by hrp2/3 RDT and/or microscopy. After informed consent, participants were enrolled in the study and 5ml of venous blood was collected.

In Ghana, samples were collected between June and July 2018 from school-aged children attending primary and junior high schools within 5 kilometers of HopeXchange Medical Centre. The eligibility for enrollment was all asymptomatic, all afebrile (axillary temperature reading below 38 degrees Celsius) students between 5 and 17 years old. Informed consent was obtained from a parent or guardian for each participant.

In Tanzania samples were collected in 2004 from Mlandizi Health Centre through passive case detection. The eligibility criteria for the study were a temperature equal to or greater than 38 °C or a fever in the past 24 hours and positive for malaria by microscopy. After informed consent, participants were enrolled in the study and 5ml of venous blood was collected.

In Burkina Faso samples were collected in 2018 during the rainy season from the Bama clinic through passive case detection. Participants were eligible if they presented with uncomplicated or severe *P. falciparum* mono or mixed species malaria infection.

The sex distribution did not differ significantly among countries (Pearson’s chi-square test, p = 0.424). In contrast, the complexity of infection (COI) differed significantly among the four study populations (Kruskal–Wallis test, P < 0.001) (Table 1).

### Overall SNP prevalence in pan-African samples

Using deep targeted amplicon sequencing, we characterized the genetic diversity of the Pfs25 gene from the 184 clinical isolates across four African countries spanning geographic locations and transmission zones. A total of 26 non-synonymous SNPs were identified across all samples. We used a very sensitive cut-off (1% variant frequency) to allow maximum SNP discovery in polygenomic infections. The Pfs25 amplicon obtained from 3D7 genomic DNA was included in the sequencing as a control for potential PCR errors to ensure SNP validity, and as expected, no SNPs were identified in the 3D7 control. The *P. falciparum* 3D7 strain served as the reference genome for all analyses.

Among the 26 SNPs identified, only 2 (L63V, V132I) had been previously reported in *Plasmodium* genomic databases^51,52^. Two additional SNPs were observed at previously described positions but with different amino acid substitutions (V143I and L63P) (Fig. 2a).

**Figure 2:**
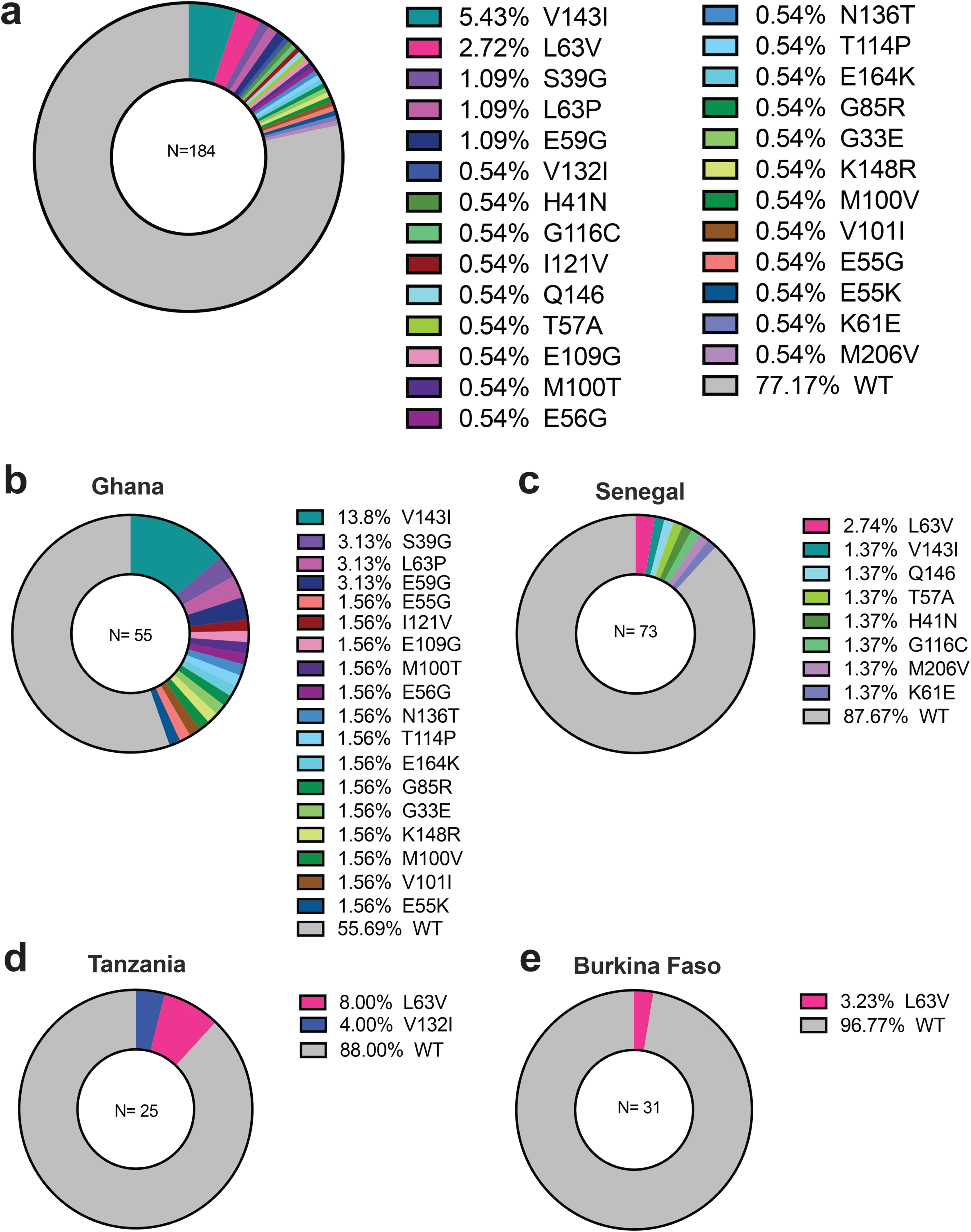
Population prevalence of novel SNPs in Pfs25. (a) The population prevalence of novel SNPs was calculated considering the total number of samples. (b-e) The population prevalence of novel SNPs was determined individually by the country of study.

Of the 26 total SNPs, 5 (L63V, V143I, S39G, L63P, E59G) were found in more than one sample, while the remaining 21 SNPs occurred as singletons, each present in only one isolate and thus classified as rare variants, at a prevalence of 0.54% each (Fig. 2a). The most prevalent allele was V143I, detected in 5.43% of samples, followed by L63V at 2.72%. Three novel SNPs (S39G, L63P, and E59G) were each detected at a prevalence of 1% from the overall set of samples we used (Fig. 2a).

### SNP Prevalence by country

Country-level analysis revealed that Ghana had the highest number of mutant alleles (44.3% of all observed SNPs), followed by Senegal (12.3%), Tanzania (12%), and Burkina Faso (3.2%). Ghana also harbored the greatest number of unique SPNs, with 17 variants found exclusively in samples from this country (Fig. 2b). Senegal followed with 7 unique SNPs (Fig. 2c). All SNPs detected in Tanzania and Burkina Faso had been previously reported, and no unique variants were identified in these countries (Fig. 2d, e). The SNP L63V was the most prevalent shared allele, occurring in samples from Tanzania (8%), Burkina Faso (3.2%) and Senegal (2.7%) (Fig. 2b, c, d, e).

A total of 18 mutant alleles were identified in Ghana: V143I, S39G, L39P, E59G, E55G, I121V, E109G, M100T, E56G, N136T, T114P, E164K, G85R, G33E, K148R, M100V, V101I, E55K. Notably, the previously reported L63V mutation exhibited an alternative amino acid substitution in our dataset: proline instead of valine (L63P). Notably, this substitution was unique to Ghana and observed at a prevalence of 3.08%. The most prevalent SNP in Ghana was V143I (13.8%), which was also found in Senegal. Two other SNPs, S39G and E59G, were unique to Ghana, each with a prevalence of 3.13% (Fig. 2c). The 14 unique variants (E55G, I121V, E109G, M100T, E56G, N136T, T114P, E164K, G85R, G33E, K148R, M100V, V101I, E55K) were classified as rare and detected at a prevalence of 1.56% each.

In Senegal, eight non-synonymous SNPs were identified in samples from Senegal: L63V, V143I, Q146* (nonsense), T57A, H41N, G116C, M206V and K61E. The most prevalent variant was L63V (2.74%), followed by V143I (1.37%), a mutation also detected in samples from Ghana. Each of the remaining six SNPs were detected at a prevalence of 1.37% (Fig. 2c). Among the 25 DNA samples from Tanzania, two previously described ^53^ SNPs were identified: L63V (8%) and V132I (4%) (Fig. 2d). No novel SNPs were observed in this cohort ^54^. In the 31 samples from Burkina Faso, a single SNP, L63V, was detected, with a prevalence of 3.23% (Fig. 2e). No unique or novel SNPs were identified in samples from Burkina Faso.

### Variant Allele Frequencies of novel SNPs within individual samples

In our dataset, the average multiplicity of infection (MOI) ranged from 2.1 to 4, depending on the country of origin (Table 1). To assess the within-sample distribution of polymorphisms, we calculated the variant allele frequency (VAF) of each SNP across all individual isolates. VAF was defined as the proportion of sequencing reads supporting the variant allele at a given position, relative to the total reads covering that site in each sample. This approach enables quantification of allele representation within complex polygenomic infections, which are common in high-transmission settings and pose challenges to haplotype resolution. We subclassified the 26 nonsynonymous SNPs identified into three categories based on their VAF: high frequency (>25%), intermediate frequency (2-25%) and low frequency (<2%), within individual samples. This distribution is visualized in Supplementary Fig.S1, where purple, green and blue bars indicate high, intermediate, and low VAFs, respectively. Among the high-frequency variants, L63V and V132I, whereas L63P and M206V represent novel mutations observed at high allele frequency within individual samples.

### Structural conformation of Pfs25 and impact of mutation

To predict the potential structural consequences of observed amino acid substitutions in Pfs25, we used PyMol v2.3.2 to thread identified SNPs onto the published crystal structure of Pfs25 in complex with the well-characterized inhibitory monoclonal antibodies mAB1269 and mAB1260 (Fig. 3a, b). Based on the threading analysis, mutations were categorized into three groups: 1) SNPs potentially impacting antibody binding at site 1 or site 2, including G131A, V132I, G116C, V143I, T57A, E59G, E56G, N136T, K148R, Q146 and G33E. These substitutions localize near or within known antibody epitopes, suggesting possible interference with antibody recognition or neutralization. 2) SNPs likely to alter Pfs25 structure, such as H41N, S39G, E109G, G85R, E55G, E55K, V101I and M206V, which may disrupt local folding or domain integrity. 3) SNPs with no predicted structural effect, such as L63V, or variants with minor or unknown structural consequences, including I121V, L63P, M100T, T114P, E164K, M100V, and K61E. Of note, the L63P substitution was unique to Ghana in our dataset and represents an alternative amino acid change at the previously reported L63 position. While substitutions involving proline may increase structural flexibility in protein regions previously considered rigid^54^, in other contexts, they may have little to no functional impact depending on the structural context. Consistent with this, structural modeling predicted only a minor effect of L63P on the overall Pfs25 conformation. For reference, we also included nine previously reported Pfs25 SNPs in the structural modeling (even if some were not detected in our current dataset: M217L, I106V, N117S, N126S, G131A, Q146E and V143A) to contextualize our observations and enable comparison ^53^ with prior studies.

**Figure 3:**
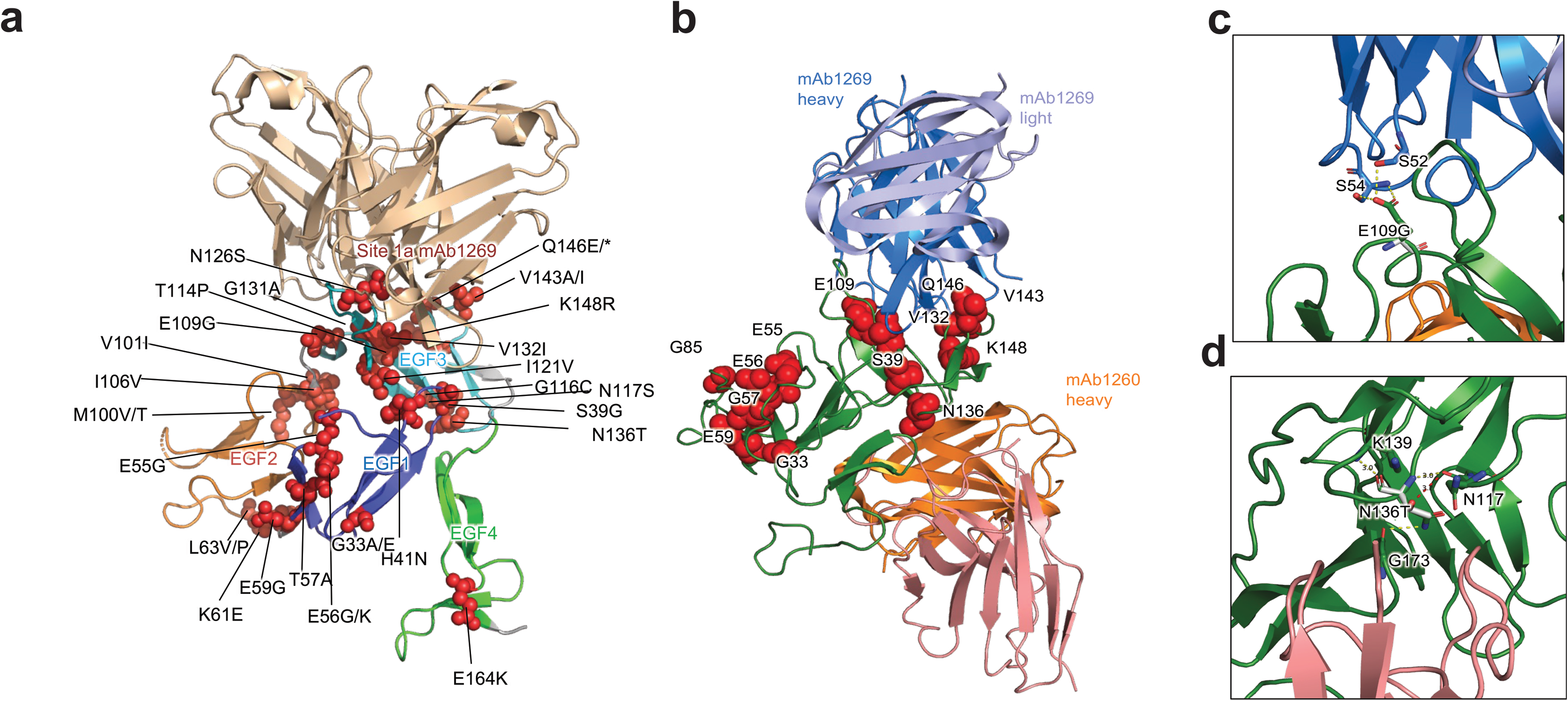
SNPs identified in Pfs25: Structure-function prediction. (a) Location of novel and known SNPs in the Pfs25 crystal structure protein in red. Pfs25 EGF domains are shown by dark blue (EGF1), orange (EGF2), light blue (EGF3), green (EGF4). The specific inhibitory monoclonal antibody mAB1269, which has an overlapping epitope with the inhibitory mAB 4B7 is shown in light beige. (b) Crystal Structure of Pfs25 (green) showing prioritized and relevant SNPs (red) and light and heavy chains of monoclonal antibodies mAB1260 (pink and orange) and mAB1269 (purple and blue) in the binding site of the protein. (c-d) Small panel highlights the predicted effect of selected SNPs.

The foldX analysis predicted that most naturally occurring Pfs25 substitutions had minimal effects on antibody interaction energies with the Site 1 antibody mAb1269 and the Site 2 antibody mAb1260 (supplementary Table S1). In contrast, several variants were predicted to destabilize Pfs25. The G116C substitution, which was selected for functional evaluation, showed predicted ΔΔG values of 3.06 and 2.86 kcal/mol in the mAb1260 and mAb1269 structural models, respectively. Other substitutions with higher predicted destabilizing effects included G33E (ΔΔG = 11.10 and 14.48 kcal/mol), G131A (ΔΔG = 0.89 and 3.36 kcal/mol), and E59G (ΔΔG = 1.59 and 2.29 kcal/mol) in the mAb1260 and mAb1269 models, respectively.

### Allelic replacement of G116C through CRISPR-Cas9 genome editing

The G116C substitution was introduced into the pfs25 locus using CRISPR-Cas9 genome editing (Fig. 4a). Following WR99210 selection, transfectant parasites were detected by microscopy 25 days post-transfection. Correct integration of the edited locus was confirmed by PCR using primer sets designed to distinguish genomic integration from episomal plasmid DNA (Fig. 4b). The observed amplicon sizes matched the expected products, and Sanger sequencing verified the presence of the G116C substitution. Limiting dilution cloning of the bulk transfectant population yielded nine positive clones, and correct integration was confirmed by PCR and sequencing (Fig. 4c, d). Two independent clones, 5B and 8E, were selected for subsequent phenotypic characterization.

**Figure 4:**
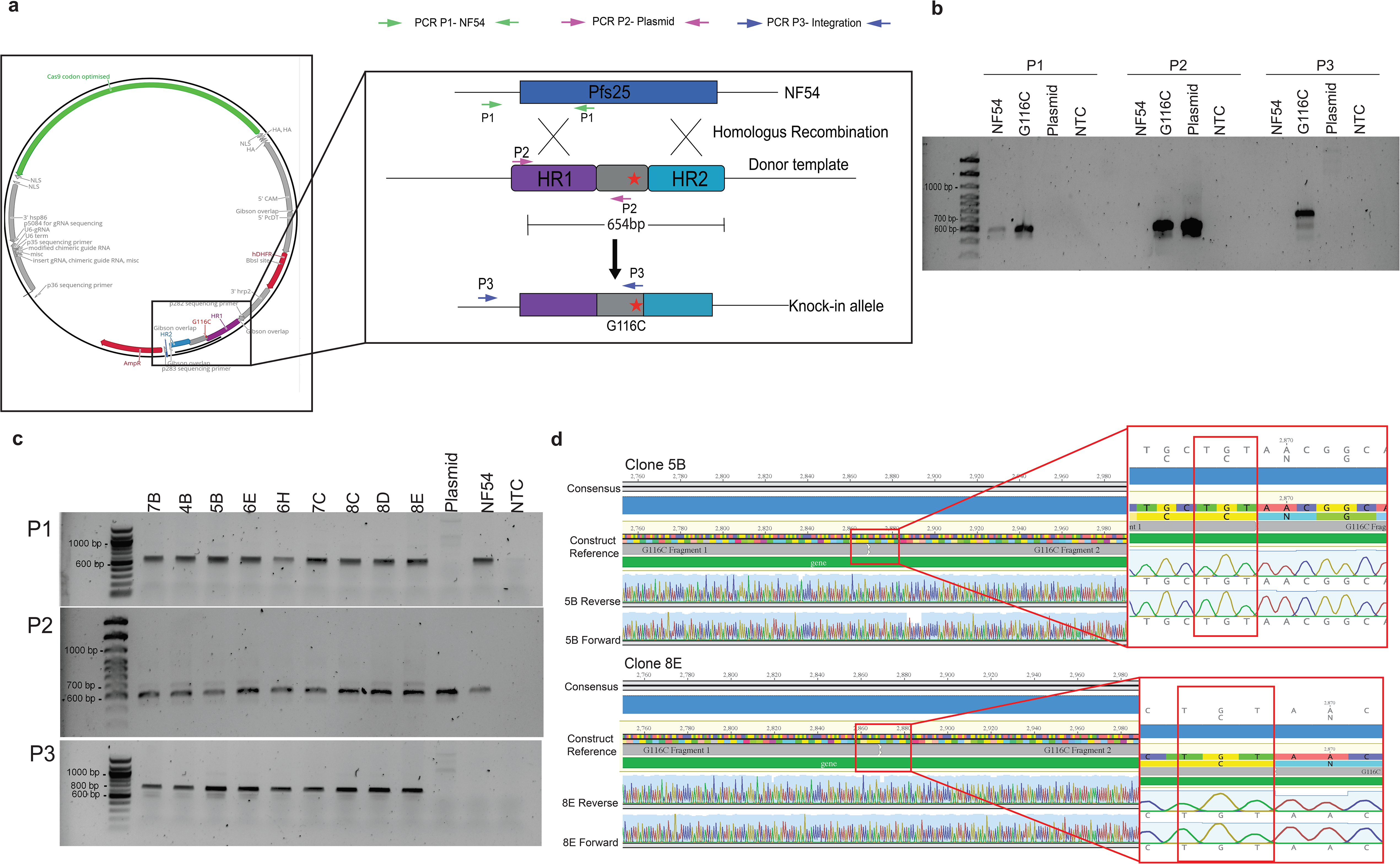
Schematic illustration and validation of the pDC2 single-vector CRISPR/Cas9 strategy. (a) pDC2 vector carrying the pfs25 gene with a point mutation introduced in the recodonized region. Donor template integration into the pfs25 locus through homologous recombination at homologous regions 1 and 2. Primers P1, P2, and P3 used for PCR-based validation are indicated. (b) Agarose gel of bulk genomic DNA PCR products from transfected parasites, confirming successful integration of the G116C mutation (P1: 646 bp; P2: 639 bp; P3: 754 bp). (c) Agarose gel of PCR products from genomic DNA of nine recovered G116C clones, all of which confirmed successful integration. All PCR products were subjected to Sanger sequencing for verification. (d) Sanger sequencing of clones 5B and 8C confirming the integration of the G116C mutation.

### The G116C mutation does not impair gametocyte development or stage progression

To determine whether the G116C mutation affected gametocyte development, NF54 and two independent G116C clones (5B and 8E) were induced to produce gametocytes in parallel. Gametocyte development was assessed morphologically by microscopic evaluation of stage distribution throughout development (Fig. 5a, c) and molecularly by quantifying the expression of stage-specific gametocyte markers using RT–qPCR (Fig. 5b, d). Gametocyte stage progression was comparable between NF54 and both G116C clones, with similar proportions of developing gametocytes observed across the different stages (Fig. 5a, c). Microscopy-based quantification of gametocyte stages showed comparable developmental trajectories across all lines (Fig. 5a, 5c). Early-stage gametocytes predominated at earlier time points, followed by a shift toward late-stage forms on the day of infection, which was corroborated by the transcriptional data. The distribution of gametocyte stages was similar between NF54 and mutant parasites, with no evidence of developmental delay or stage-specific accumulation. Together, these results indicate that the G116C mutation does not affect gametocyte development or maturation. Expression of PfHISTa, Pf48/45, and Pfs25 were measured during gametocyte development by RT-qPCR, using ubiquitin-conjugating enzyme (UCE) as the reference gene (Fig. 5b, d). The early gametocyte marker PfHISTa (PF14_0748), showed positive ΔCt values, exhibiting lower relative expression compared with the housekeeping gene ubiquitin-conjugating enzyme. Pf48/45 displayed a peak in expression at mid-late stages, with lower ΔCt values around days 8–10 (Fig. 5b, d). Pfs25, a late-stage gametocyte marker, exhibited progressively lower ΔCt values at later time points consistent with stage IV–V gametocyte maturation (Fig. 5b, d). Error bars represent mean ± SD of two biological replicates, and similar trends were observed across NF54 and the two mutant lines 5B and 8E. To confirm that the edited parasites retained the mutation, parasites were collected on the day of infection prior to mosquito feeding. Genomic DNA was extracted and subjected to sequencing, which confirmed that the gametocytes used for infection had the targeted mutation (Fig. 5e).

**Figure 5:**
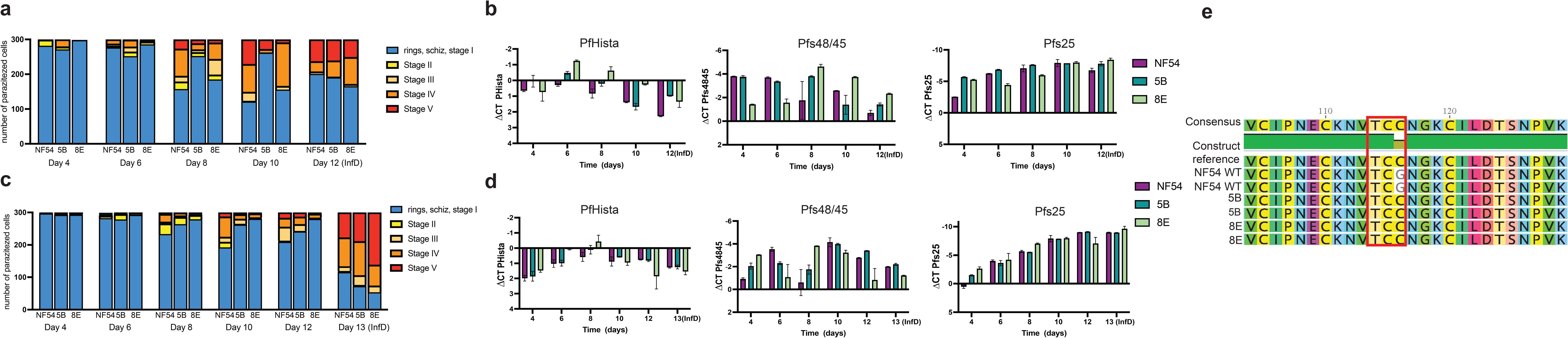
Characterization of gametocyte conversion in the mutant lines of *P. falciparum* 5B and 8E compared to NF54. (a-c) In two independent experiments, gametocytes were induced and samples were collected every other day starting from day 4 until the day of infection (InfD). A total of 300 parasitized cells were counted, and gametocytes were identified based on the morphology into the five known stages (b-d) The two independent experiments showing the stage specific expression markers of gametocyte along the induction. ΔCt values were calculated as (Ct target gene – Ct reference) and were plotted on an inverted y-axis, where more negative values indicate higher transcript abundance. Panels b and d represent independent qPCR experiments, both showing consistent temporal expression patterns PfHista is specific to stages I and II, PFs48/47 to stages III and IV and Pfs25 to stages IV and V. The normalized expression doesn’t show a difference between NF54 and the clones 5B and 8E during the induction process. The bars are SD between two biological replicates. (e) Protein sequence alignment of NF54 WT, 5B, and 8E parasite lines based on sequencing performed on the day of mosquito infection, confirming the presence of the G116C mutation (boxed) in the mutant clones compared to NF54 WT.

### The G116C mutation does not affect parasite infection of the mosquito midgut

To evaluate whether the G116C mutants produce functional gametocytes capable of infecting mosquitoes, we performed standard membrane feeding assays (SMFA) using WT NF54 as a control. For all groups, cultures were adjusted to the same final gametocytemia and exflagellation rates before feeding. The NF54 parasites established robust infections, with a median of 2 oocysts per mosquito and a prevalence of 65%. In contrast, mutant clones 5B and 8E showed severely impaired infectivity, with prevalences of 33% and 20%, respectively, and median oocyst counts of 0. A few mosquitoes had low numbers of midgut oocysts, but overall infection intensity was significantly reduced (p= 0.0256 and 0.0129 for 5B and 8E, respectively) compared with the control (Fig. 6a). Exflagellation rates on the day of infection, calculated as the average number of exflagellation per field across 10-11 fields, were comparable among all groups (NF54 [5.2]; 5B [4.3]; 8E [9.6]), with clone 8E showing the highest exflagellation activity. The infection prevalence was compared by Fisher’s exact test, showing no significant difference among groups.

**Figure 6:**
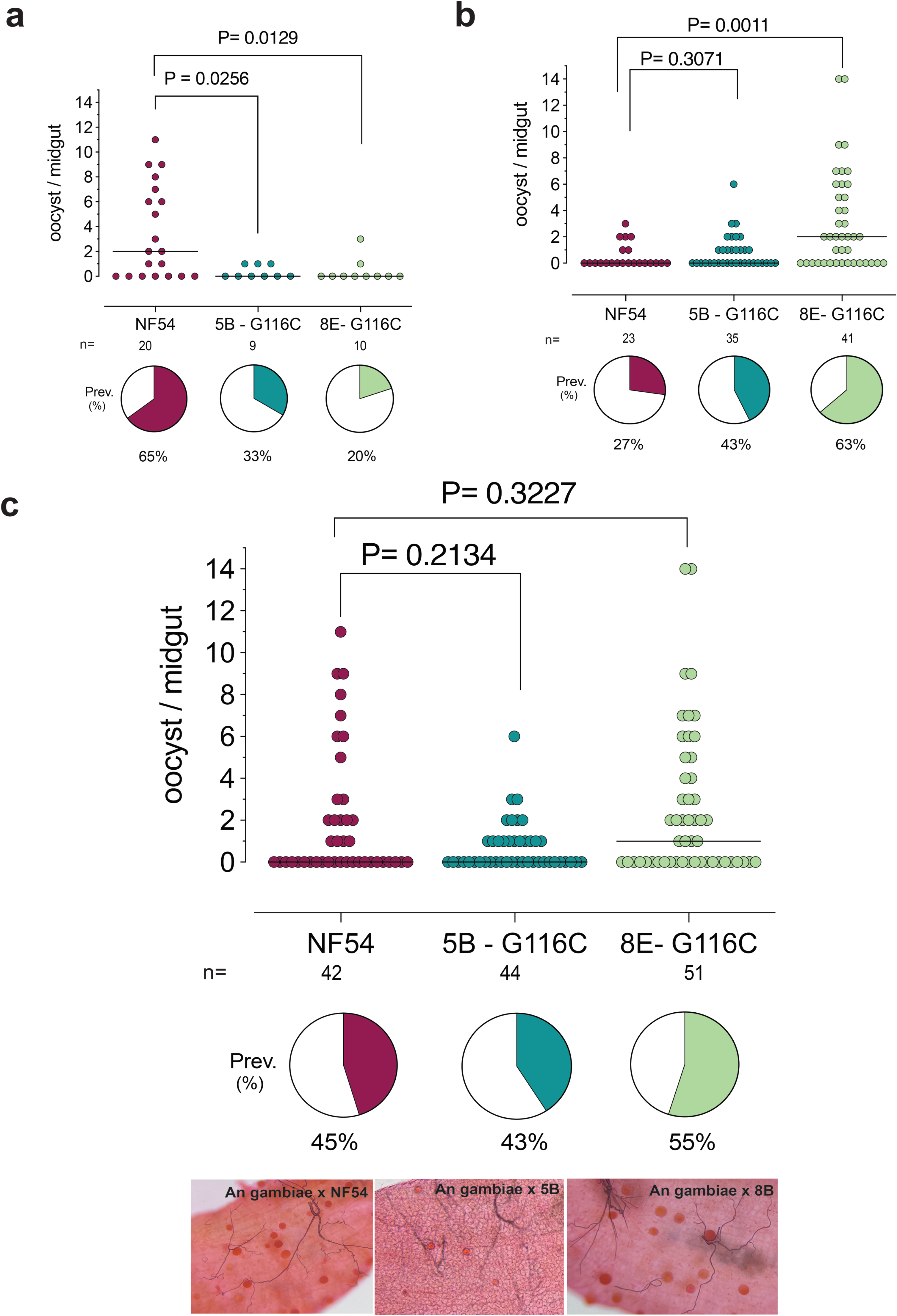
Infectivity of G116C mutant parasites in *An. gambiae*. (a) Prevalence of infection (pie charts) and oocyst counts per midgut, measured 8–10 days post-infection with NF54 -WT, and mutants 5B and 8E parasites from a single SMFA. Each dot indicates the oocyst count on an individual mosquito midgut, and the black horizontal line shows the median of oocysts burden. Medians were compared using the Mann–Whitney test (p = 0.0256 for clone 5B, p = 0.0129 for clone 8E vs NF54), and infection prevalence was compared by Fisher’s exact test, showing no significant difference among groups. (b) Independent repeat experiment showing oocyst intensity for the same parasite lines. The Mann-Whitney test showed no significant difference between NF54 and clone 5B (P= 0.3071) and a statistical difference between NF54 and clone 8E (P=0.0011), the chi-square test showed a significant difference among groups. Pairwise comparisons with Bonferroni correction (P = 0.0167) revealed a significant difference only between group NF54 and clone 8E (P = 0.0062) (c) Combined analysis of both experiments showed no statistical difference in the median of oocysts between the groups (P = 0.2134 and 0.3227, respectively, for 5B and 8E). Prevalence of infection also showed no statistical difference according to the chi-square test (P = 0.3728). Representative infected midguts for each parasite line are shown below.

In a second independent experiment (Fig. 6b), infection prevalence was higher in the mutant groups. Additionally, only clone 8E showed a significant increase in infection intensity compared to NF54 (P = 0.0011), while 5B remained comparable to the WT (not significant). Consistent with this, 8E exhibited a greater proportion of infected mosquitoes (63%) compared to NF54 (27%) and 5B (43%), indicating variability in transmission efficiency across experiments. A chi-square test confirmed a significant difference among groups, and pairwise comparisons with Bonferroni correction (α = 0.0167) revealed a significant difference only between NF54 and clone 8E (P = 0.0062).

To assess the overall effect across experiments, data from both assays were combined (Fig. 6c). In the pooled analysis, no statistically significant differences in infection intensity were observed among NF54, 5B, and 8E. Infection prevalence was also comparable across groups (45%, 43%, and 55%, respectively) with no statistical difference detected by chi-square test (P = 0.3728). Together, these findings indicate that G116C mutant parasites retain the ability to infect mosquitoes at levels similar to WT when data are considered collectively, suggesting that this mutation does not significantly impair transmission. Representative midgut images are shown in figure 6c, where oocysts were readily observed in mosquitoes infected with WT NF54 and both G116C mutant clones, demonstrating successful parasite development in the mosquito midgut.

## Discussion

As with many other transmission-blocking antigens, the pfs25 gene is highly conserved gene relative to many other antigenic targets in the P. falciparum genome^55^. This conservation is due to the specific kinetics of protein expression: Pfs25 is expressed in the post fertilization stages^56^ and as such is not subject to the selective pressure of the human immune response^38,57,58^. Both dN/dS ratios and Tagima’s D values are much lower than other major vaccine candidate antigens such as AMA1 and MSP1 in the blood stages and pre-erythrocytic antigen CSP^55^. The genetic diversity that is present in Pfs25 and other sexual antigens expressed in the post-fertilization stages are likely driven by factors related to adaptation to and transmission through the mosquito vector^59^.

The present study characterized the genetic diversity of the pfs25 gene in 4 different African countries with unique malaria epidemiological contexts: Burkina Faso, Ghana, Senegal, and Tanzania. Previous studies demonstrated that pfs25 is a conserved gene with maximally 30 non-synonymous SNPs^53,57,58,60^. Our analysis of the Pf-HaploAtlas^52^ dataset confirmed the predominance of the wild-type pfs25 haplotype and identified only a limited number of recurrent mutant haplotypes. Among the variants detected in our study, L63V and V132I were also present in the global dataset. However, their geographic distribution differed, as these variants were not reported in the same countries in which we identified them (L63V in Senegal, Burkina Faso and Tanzania, and V132I in Tanzania). The remaining variants detected in our study were not represented in the current Pf-HaploAtlas^52^, suggesting that they are either rare, geographically localized, or remain underrepresented in available genomic datasets. Our results revealed 26 non-synonymous SNPs across all sites, and of these 24 were novel SNPs, including variants that might be responsible for the immune evasion of transmission blocking antibodies. Most of the SNPs discovered in this study were in domain three (D3) of Pfs25, which is the most potent inhibitor site of the protein and contains the binding sites for the transmission blocking monoclonal antibodies, including 1D2, 32F81 and 4B7 which is used in the standard membrane feeding assays for *P. falciparum*^61–64^. The high variability of non-synonymous SNPs observed in our dataset, particularly concentrated in D3 of Pfs25, is consistent with previous reports on Pfs25 genetic diversity^57,58^.

To investigate the functional implications of the SNPs identified in this study on parasite infection and immune evasion, we first threaded each SNP into the pfs25 crystal structure. This analysis revealed that several mutations may impact the binding site between the pfs25 protein and the transmission-blocking antibodies. Among these, the V132I has been predicted as one of the variants that may have that antibody impact. This same SNP has also been reported in other studies in Kenya^57^, Cameron, Congo, Malawi, Tanzania^60^. The following set of polymorphic residues (L63V, V132I, Q146E, V143A, G131A, N117S, G116C, V143I) are in the epitope of potent Pfs25 antibodies such as mab4B7^63^, mab2544^65^, mab1269^61^, mab1245^61^. The impact of these residues of Pfs25 on the binding affinities of the antibodies or SMFA assays has never been characterized, even for previously reported residues (L63V, V132I).

These findings highlight the importance of evaluating SNPs for their potential roles in immune evasion and transmission. While constructs were generated for multiple prioritized SNPs, only functional data for G116C are reported here, demonstrating successful transfection into *P. falciparum* and feasibility for functional studies. Moreover, the recovery timeline supports its use for gametocyte induction, as optimal conversion rates are typically maintained for 2–3 months after thawing from liquid nitrogen^66^. These results provide a foundation for assessing how specific SNPs may influence parasite development and transmission potential.

Additionally, the SNP V143I identified here is novel. While the codon at position 143 has previously been reported to harbor nonsynonymous mutations, these involved substitutions to different amino acids V143A^58,67–69^ and V143G^57^. The repeated emergence of distinct amino acid changes at the same codon suggests that this site may represent a mutational hotspot under selective pressure (mosquito immune response, parasite competition during co-infection in the mosquito midgut), with potential functional consequences for protein structure or immune recognition. The identification of V143I therefore highlights the need for further functional studies to determine whether substitutions at this position confer an advantage in parasite survival, transmission, or immune evasion.

Together, our findings demonstrate that the G116C substitution in Pfs25 does not compromise early sexual development, as evidenced by comparable gametocytemia and exflagellation rates between mutant clones and WT NF54 line. These results suggest that gametocyte maturation and male gamete formation remain intact despite the mutation. Notably, when oocyst data from independent experiments were combined, no significant difference in the overall number of oocysts was observed, indicating that the allelic replacement does not substantially affect parasite establishment within the mosquito midgut at the population level. Similarly, differences in oocyst prevalence and media counts were not consistently reproduced across replicates. Collectively, these findings suggest that the G116C substitution does not have a measurable impact on parasite transmission to *An. gambiae* G3 and that the variability observed across experiments likely reflects the inherent variability of the SMFA as previously reported^70,71^.

Structural analyses have shown that proper folding of the EGF-like domains in Pfs25 is essential for maintaining its function and stability^61,72^. Although in silico modeling (supplementary table S1) predicted that the G116C substitution in Pfs25 could moderately destabilize the protein, ΔΔG = + 2.86 kcal/mol, combined mosquito infection experiments demonstrated that mutant clones 5B and 8E were capable of establishing infections at levels comparable to, and in some replicates higher than WT NF54. These results indicate that the predicted thermodynamic destabilization does not result in a strong functional defect under physiological conditions encountered during mosquito infection. Pfs25 is expressed on the surface of the zygotes and ookinetes and it has been implicated in ookinete survival in the protease rich midgut lumen, penetration of the midgut epithelium and oocyst maturation, making it a key post-fertilization antigen targeted by transmission-blocking strategies^20,73,74^. Our functional data therefore suggest that at least some naturally occurring Pfs25 variants may be tolerated without measurable effects in parasite transmission.

Taken together, our findings broaden the current understanding of Pfs25 genetic diversity and its implications for malaria transmission-blocking vaccine development. By identifying 26 non-synonymous SNPs, including 24 novel variants across diverse geographic regions, we provide new insights into the genetic variability of *P. falciparum* in malaria-endemic areas. Notably, the majority of SNPs identified were concentrated in domain D3, which is located in EGF domain III and corresponds to the epitope for the Pfs25 specific transmission blocking mAb, 4B7^26,75^, which emphasizes the need for further investigation into its role in immune evasion and transmission dynamics. These findings build on existing evidence of Pfs25 conservation while highlighting subtle variations that could influence the efficacy of transmission-blocking antibodies.

### Limitations of the study

Despite its contributions, this study has limitations that should be considered when interpreting our findings. Although comparison with the MalariaGEN Pf8^51^ dataset through Pf-HaploAtlas^52^ provided broader geographic context and further supported the high conservation of pfs25, our primary analyses included samples from only four African countries and may not fully capture the global diversity of *P. falciparum* populations or regional variations in transmission dynamics. However, the deep targeted amplicon sequencing approach used in this study provides greater sensitivity for detecting low frequency variants than whole genome sequencing, allowing the identification of rare mutations that may be missed by large scale genomic surveillance datasets. Second, the number of samples per country was relatively modest, limiting the ability to detect extremely rare variants or fully characterize population-level prevalence. Third, while constructs were generated for several SNPs, functional characterization was performed only for G116C, meaning the effects of other variants on parasite biology, immune evasion, or transmission remain unknown. Future studies will expand these analyses to additional naturally occurring Pfs25 variants and evaluate the impact of these variants on antibody binding and transmission blocking activity to determine whether these polymorphisms can influence vaccine efficacy.

Malaria genomic epidemiology provides valuable insights for surveillance and informing strategies to improve TBVs. Together, our findings underscore the complexity of *P. falciparum* populations in high-transmission regions for malaria and highlight the importance of integrating within-host and population-level measures of diversity when evaluating variant dynamics and anticipating potential challenges for transmission-blocking interventions. Incorporating genomic insights into vaccine development pipelines will be critical to overcoming the obstacles posed by parasite genetic diversity and achieving malaria elimination.

## Data Availability

The raw sequencing data generated in this study have been deposited in the NCBI Sequence Read Archive (SRA) under accession number PRJNA1204390. All other relevant data supporting the findings of this study are available within the article and its supplementary information files or from the corresponding author upon reasonable request.

https://www.ncbi.nlm.nih.gov/sra/?term=PRJNA1204390

## Acknowledgements

This work was supported by an International Research Scientist Development Award (K01 TW010496) from the Fogarty International Center and a R01 (R01 AI168238) from the National Institute of Allergy and Infectious Diseases, of the National Institutes of Health to A.K.B. and by CTSA Grant Number UL1 TR001862 from NCATS/NIH to A.O. Additional support was provided by the Ambrose Monell Foundation and the Fulbright Program. The funders had no role in study design, data collection and analysis, decision to publish, or preparation of the manuscript.

We are grateful to Prof. Flaminia Catteruccia for providing the G3 eggs for our Anopheles gambiae colony. Strain NF54 (Patient Line E), MRA-1000, was obtained through BEI Resources, NIAID, NIH: Plasmodium falciparum, and was contributed by Megan G. Dowler.

We thank the Yale Center for Genome Analysis (YCGA) for providing next-generation sequencing services for this study. Research reported in this publication was supported by the National Institute of General Medical Sciences of the National Institutes of Health under Award Number 1S10OD030363-01A1. We are grateful to all the participants who participated in the study and to their communities.

## Author contributions

A.K.B. conceived the study. M.C., S.P., B.D.F, R.K.D, J.B.O., F.A.S, Z.P., I.V-W. supervised research studies that contributed samples. L.G.T, A.B., R.L., M.N.P., K.M., A.J.M., F.D., S.D.S., B.D.S, A.M., C.M., B.N., T.B., N.O. collected and processed samples for genomics studies. A.O., A.C., L.H. performed the experiments. Y.G., Z.S., S.D.P., and L.S. performed structure modeling. A.K.B., A.C., N.F., E.M.N., and A.O analyzed the data. A.K.B and A.O. wrote the manuscript. All authors reviewed the manuscript.

## Supplemental information

Document S1. Supplemental Figure S1 and Table S1 and S2

