## Supplemental Files for "Identification of genetic variants in Pfs25 and functional evaluation in mosquito infection"

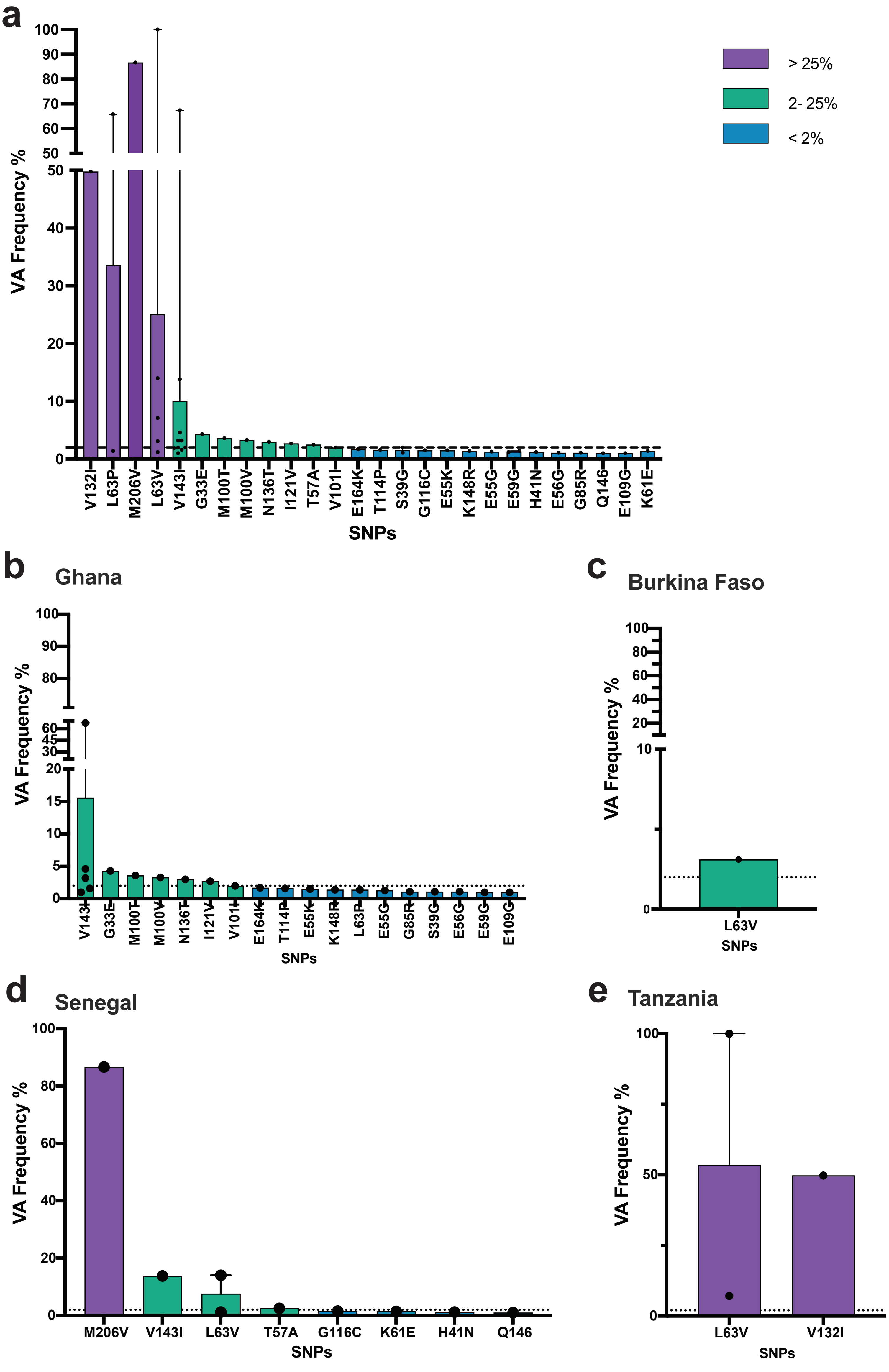


**Fig S1**: (**a**) Overall VA frequency of Pfs25 SNPs. The frequency of novel SNPs was categorized as high above 25%(purple), intermediate between 2-25% (green), and low below 2%(blue). (**b-e**) Frequency of SNPs determined by the country of study.

**Table S1**. FoldX-predicted effects of Pfs25 non-synonymous SNPs on antibody binding.

| ID antibody | Mutations | Interaction_energy_change (kcal/mol)[1] | Pfs25_stability_change (kcal/mol)[2] |
|---|---|---|---|
| mAb1260 | V132I | 0.00 | -0.38 |
| mAb1260 | V143I | 0.00 | 0.20 |
| mAb1260 | S39G | 0.01 | **0.86** |
| mAb1260 | T57A | 0.00 | **0.70** |
| mAb1260 | E109G | 0.00 | -0.20 |
| mAb1260 | E59G | 0.00 | **1.59** |
| mAb1260 | E56G | -0.01 | -0.13 |
| mAb1260 | N136T | **0.66** | -0.19 |
| mAb1260 | G85R | 0.00 | -0.46 |
| mAb1260 | K148R | 0.04 | 0.47 |
| mAb1260 | E55G | -0.01 | **0.58** |
| mAb1260 | E55K | -0.03 | 0.22 |
| mAb1260 | G33E | 0.06 | **11.10** |
| mAb1260 | L63V | 0.00 | *0.61* |
| mAb1260 | G131A | 0.00 | **0.89** |
| mAb1260 | H41N | 0.35 | *-0.83* |
| mAb1260 | G116C | 0.02 | **3.06** |
| mAb1260 | I121V | 0.00 | **0.55** |
| mAb1260 | L63P | 0.00 | 0.06 |
| mAb1260 | M100T | 0.00 | 0.39 |
| mAb1260 | T114P | -0.11 | **1.81** |
| mAb1260 | M100V | 0.03 | -0.16 |
| mAb1260 | Q146* | NA | NA |
| mAb1260 | V101I | -0.20 | -0.19 |
| mAb1260 | K61E | **0.59** | **0.92** |
| mAb1260 | M206V | NA | NA |
| mAb1269 | V132I | 0.26 | **0.87** |
| mAb1269 | V143I | -0.24 | -0.06 |
| mAb1269 | S39G | 0.04 | **1.03** |
| mAb1269 | T57A | 0.00 | *0.62* |
| mAb1269 | E109G | **1.09** | 0.10 |
| mAb1269 | E59G | 0.00 | **2.29** |
| mAb1269 | E56G | 0.00 | 0.43 |
| mAb1269 | N136T | -0.04 | -0.23 |
| mAb1269 | G85R | 0.00 | **2.02** |
| mAb1269 | K148R | 0.40 | *-0.79* |
| mAb1269 | E55G | 0.00 | **0.96** |
| mAb1269 | E55K | -0.02 | 0.05 |
| mAb1269 | G33E | 0.00 | **14.48** |
| mAb1269 | L63V | 0.00 | 0.48 |
| mAb1269 | G131A | 0.01 | **3.36** |
| mAb1269 | H41N | -0.15 | **0.59** |
| mAb1269 | G116C | 0.04 | **2.86** |
| mAb1269 | I121V | 0.20 | **0.72** |
| mAb1269 | L63P | 0.00 | *-0.62* |
| mAb1269 | M100T | 0.00 | **1.26** |
| mAb1269 | T114P | 0.37 | -0.39 |
| mAb1269 | M100V | 0.00 | **1.18** |
| mAb1269 | Q146* | NA | NA |
| mAb1269 | V101I | -0.03 | *-0.66* |
| mAb1269 | K61E | 0.00 | **1.16** |
| mAb1269 | M206V | NA | NA |

(1) predicted interaction energy change with the antibody (ΔΔG, kcal/mol), and (2) predicted effect on Pfs25 including interaction energy and protein stability changes (ΔΔG, kcal/mol). Positive interaction energy changes (> +0.5 kcal/mol) indicate weaker antibody binding (potential escape), while negative values indicate stronger binding; values within ±0.5 kcal/mol are considered near-neutral. Protein stability changes reflect the effect of mutations on *pfs25* folding: positive values (> +0.5 kcal/mol) indicate destabilization, and negative values indicate stabilization. Variants with very high destabilization (> +2 kcal/mol) may be biologically non-viable regardless of antibody interaction. NA denotes mutations where FoldX did not return values (e.g., stop codons or residues not modeled in the complex). Color coding highlights predicted effects: bold = destabilizing/reduced binding, italic and underline = stabilizing/enhanced binding, gray = near-neutral.

**Table S2**. Primer pairs used in PCR and in qRT-PCR assays.

| Primer | Sequence 5'-3' | Prod. (bp) |
|---|---|---|
| Ubiquitin conjugating enzyme (all stages) F | GGTGTTAGTGGCTCACCAATAGGA | qPCR |
| Ubiquitin conjugating enzyme (all stages) R | GTACCACCTTCCCATGGAGTATCA | |
| PfHISTA (Gam1-2) F | ATTCAAGGGTAGTTCCTAGAGCAGTGTGG | qPCR |
| PfHISTA (Gam1-2) R | AGCACTCGTAATTCTAACACTGGG | |
| Pfs48/45 (gams 3-4) F | GTAAGCCTAGCTCTTTGA ATAGTGA | qPCR |
| Pfs48/45 (gams 3-4) R | GACCTACGTTCACGCATATCTGGCT | |
| Pfs25 (gams 4-5) F | GGAGAAACCAAATGCTCATTAAAATG | qPCR |
| Pfs25 (gams 4-5) R | GCAGTACATATAGAGCTTTCATTATC | |
| B1 overlap F Pfs25 | GAGGTACCGAGCTCGAATTCAATACATATTATTAATTTTA | 531 |
| Pfs25 WT synthetic Rv | GTCGCACTTCAGAACTTTTTCTTCAC | |
| A4 overlap R Pfs25 | CGAAAAGTGCCACCTGACGTCTAATTTACCATTTAAAATA | 294 |
| Pfs25 to synthetic F | CAACGTGCAGGACCAGAATAAATGTTCAAAAG | |
| Pfs25 FP1 | CTTATAGAGTTTGGCTAGATATATAAG | 646 |
| Pfs25 RP1 | GTCGCACTTCAGAACTTTTTCTTCAC | |
| Pfs25 - FP2 | GAGGTACCGAGCTCGAATTCAATACATATTATTAATTTTA | 639 |
| Pfs25 - RP2 | GTTCACCATGTCGTAGCC | |
| Pfs25 - FP3 | CTTATAGAGTTTGGCTAGATATATAAG | 754 |
| Pfs25 - RP3 | GTTCACCATGTCGTAGCC | |
